# Deep learning for quantitative MRI brain tumor analysis

**DOI:** 10.1101/2023.03.21.23287514

**Authors:** Iulian Emil Tampu, Neda Haj-Hosseini, Ida Blystad, Anders Eklund

## Abstract

The infiltrative nature of malignant gliomas results in active tumor spreading into the peritumoral edema, which is not visible in conventional magnetic resonance imaging (cMRI) even after contrast injection. MR relaxometry (qMRI) measures relaxation rates dependent on tissue properties, and can offer additional contrast mechanisms to highlight the non-enhancing infiltrative tumor. The aim of this study is to investigate if qMRI data provides additional information compared to cMRI sequences (T1w, T1wGd, T2w, FLAIR), when considering deep learning-based brain tumor (1) detection and (2) segmentation. A total of 23 patients with histologically confirmed malignant glioma were retrospectively included in the study. Quantitative MR imaging was used to obtain R_1_ (1/T1), R_2_ (1/T2) and proton density maps pre- and post-gadolinium contrast injection. Conventional MR imaging was also performed. A 2D CNN detection model and a 2D U-Net were trained on transversal slices (n=528) using either cMRI or a combination of qMRI pre- and post-contrast data for tumor detection and segmentation, respectively. Moreover, trends in quantitative R_1_ and R_2_ rates of regions identified as relevant for tumor detection by model explainability methods were qualitatively analyzed. Tumor detection and segmentation performance for models trained with a combination of qMRI pre- and post-contrast was the highest (detection MCC=0.72, segmentation Dice=0.90), however, improvements were not statistically significant compared to cMRI (detection MCC=0.67, segmentation Dice=0.90). The analysis of the relaxation rates of the relevant regions identified using model explainability methods showed no differences between models trained on cMRI or qMRI. Relevant regions which fell outside the annotation showed changes in relaxation rates after contrast injection similar to those within the annotation, when looking at majority of the individual cases. A similar trend could not be seen when looking at relaxation trends over all the dataset. In conclusion, models trained on qMRI data obtain similar performance to those trained on cMRI data, with the advantage of quantitatively measuring brain tissue properties within the scan time (11.8 minutes for qMRI with and without contrast, and 12.2 minutes for cMRI). Moreover, when considering individual patients, regions identified by model explainability methods as relevant for tumor detection outside the manual annotation of the tumor showed changes in quantitative relaxation rates after contrast injection similar to regions within the annotation, suggestive of infiltrative tumor in the peritumoral edema.

## 1. Introduction

Malignant gliomas are tumors of the central nervous system with high recurrence and high mortality rates, as well as poor prognosis (Davis, 2016). Magnetic resonance (MR) images are essential for the diagnosis and treatment follow-up of malignant gliomas and brain tumors in general, with T1-weighted pre- and post-gadolinium contrast (T1w and T1wGd), T2-weighted (T2w), fluid-attenuated inversion recovery (T2wFLAIR), perfusion and diffusion-weighted images routinely acquired (Juratli et al., 2019). Radiologists use these images for first diagnosis and to delineate the tumor structure to balance the extent of the treatment with the possible collateral effects. However, the infiltrative nature of malignant gliomas poses a great challenge in delineating the tumor boundary. In fact, not all the active and infiltrative tumor regions are enhanced and visible in the routinely acquired MR images (Konukoglu et al., 2010), which can lead to tumor regrowth if not considered during treatment. Targeting the whole tumor region during treatment has shown to positively impact disease progression as well as mortality rate (Brown et al., 2016). To aid clinicians to safely and accurately understand the extent of tumor treatment region, new non-invasive imaging and analysis methods are needed. In comparison to conventional MRI sequences (T1w, T1wGd, T2w, FLAIR) hereafter cMRI, where contrast between tissues is obtained by tuning the acquisition sequence parameters, quantitative imaging approaches using MRI measure tissue properties related to the aqueous composition and functionality of the tissue (Warntjes et al., 2008; Gurney-Champion et al., 2020). Among the proposed quantitative MR imaging sequences, diffusion-weighted MRI (Maier et al., 2010), perfusion MRI (Cha, 2004) and MR relaxometry (Hattingen et al., 2015), are the most common approaches used today. By measuring tissue properties, quantitative MRI has the possibility of complementing the morphological analysis currently performed by clinicians on conventional images with quantitative information from normal and abnormal tissue. In addition, it could also aid early diagnosis since deviations from normal tissue values could be detected prior to the manifestation of visible morphological changes (Keenan et al., 2019, 2022). In particular, several studies have shown the potential of MR relaxometry (qMRI) in clinical applications (Cashmore et al., 2021). However, the limited knowledge of the relaxation rates of healthy and diseased brain tissue hinders its mainstream application in the clinical workflow. In the context of brain tumors, quantitative relaxation maps of patients diagnosed with malignant gliomas have been used to show that tumor-like T1 and T2 relaxation times extend in the peritumoral edema beyond the contrast enhanced tumor region visible in cMR images (Blystad et al., 2017). These findings suggest MRI relaxometry as a potential quantification method of the non-enhancing infiltrative tumor, which could be used to generate new ways of looking at the active tumor region. This could help radiologists to better delineate the tumor, as well as to enabling early detection of tumor growth with no or subtle morphological changes in conventional images.

In the context of medical image analysis, deep learning methods can be trained to solve, among others, segmentation and detection tasks (Lee et al., 2017). Compared to traditional imaging processing approaches where features are manually engineered, deep learning methods automatically learn task-specific features from the data in a data-driven optimization fashion. Deep learning methods have been successfully implemented for the analysis of cMR images of gliomas, where convolutional neural networks (CNN) have been trained to perform tumor segmentation to reduce the burden of manual annotation for radiologists, and providing an objective method for tumor boundary delineation (Gryska et al., 2021). Moreover, such methods have also been implemented on quantitative MR imaging protocols (Gurney-Champion et al., 2022). For example, perfusion MR data alone was used as model input for automatic tumor segmentation by (Jeong et al., 2020) showing a Dice score up to 0.9, while (Rahmat et al., 2020) investigated the use of diffusion-weighted MR data during model training, showing that a good tumor segmentation could be achieved only when spatial context information from T1wGd and FLAIR was combined with diffusion-derived metrics (Dice score of 0.82). Given the success of deep learning in analyzing conventional (Bakas et al., 2018) and quantitative MR data of gliomas, such methods could be used to learn tumor-specific features from the MRI relaxometry data. These methods can then be employed to generate new ways of looking at the tumor that may help identify the non-enhancing infiltrative tumor.

Thus, the aim of this work was to investigate if MRI relaxometry data provides additional information compared to the cMR image sequences (T1w, T1wGd, T2w, T2wFLAIR) when considering deep learning-based brain tumor (1) detection and (2) segmentation. Moreover, using model explainability methods, the regions identified as relevant for tumor detection by models trained on different input configurations were qualitatively investigated. This method could uncover radiological biomarkers previously invisible in the conventional MRI, which could be useful for tumor treatment planning and evaluation.

## 2. Material and methods

### 2.1. Dataset description

Twenty-three patients with typical radiological findings suggestive of a high-grade malignant glioma were retrospectively included in a study from 2013 to 2016 and examined with MRI before surgery. Mean age at inclusion was 61 years (range 34-82), six patients were females. Ethical approval was obtained from the regional ethical board of Linköping, Sweden (decision number 2011 / 406-31) and informed written consent was obtained from all patients. For all the patients, axial T1w, T1wGd, T2w and T2wFLAIR images (referred to as cMRI in this paper) were acquired on a 3-tesla MR scanner (750, GE Medical Systems, Milwaukee, Wisconsin) using a 32-channel phased array head coil according to the clinical protocol for brain tumor investigation specified by Linköping University hospital. The total scan time for the cMRI data per patient was 12.2 minutes. A high resolution 1 mm isotropic T1wGd volume (BRAVO volume) was also acquired and used to manually annotate the tumor. In addition, qMRI pre- and post-gadolinium contrast images were acquired using a multi-slice, multi-echo and multi-saturation delay sequence for simultaneous measurement of R_1_ (1/T1), R_2_ (1/T2), and proton density (PD) (Warntjes et al., 2008). The qMRI data was processed using the software SyMRI (version 8, developed by SyntheticMR AB, Linköping Sweden) to obtain volumetric maps of T1- and T2-relaxations, as well as proton density. The total scan time for the qMRI data was 11.8 minutes per patient. A detailed description of the acquisition protocols for each image sequence is provided in Appendix A. Two of the 23 subjects were excluded due to incomplete data (missing BRAVO volume). In total, ten image sequences were available for each of the remaining 21 subjects: four cMRI (axial T1w, T1wGd, T2w and T2wFLAIR), three qMRI pre-contrast (T1, T2 and PD) and three qMRI post-contrast (T1Gd, T2Gd and PDGd). For each subject, the tumor core (necrotic region ⋃ enhanced region) was manually annotated by an expert neuroradiologist on the BRAVO volume and then registered to the axial T1wGd volume (see section 2.2). Although there are many open access datasets containing MR images of brain tumors (Adem et al., 2022; Magadza and Viriri, 2021), we are not aware of a dataset that contains both qMRI and cMRI data. Compared to the popular BraTS dataset (Menze et al., 2009), our dataset contains the same MR images, as well as qMRI before and after gadolinium contrast. The clinical protocol used at Linköping University Hospital also contains diffusion and perfusion imaging, but those images were not included in this investigation to keep the cMRI data similar to that of BraTS. Moreover, the manual annotations available in this study do not account for the edema region, and do not have separate labels for the necrotic and tumor enhancing region.

### 2.2. Data preprocessing

All DICOM files were first converted to NIFTI volumes using the AFNI software (Cox, 1996). To correct for head motion between the different scans of each patient, a rigid body registration was performed between the axial T1wGd volume and all other volumes. No registration was performed between the different patients. For the qMRI data, the registration of the pre- and post-contrast volumes was performed independently. First the T1 and T1Gd volumes were registered to the axial T1wGd volume, and the obtained registration parameters were then applied to pre- and post-contrast qMRI volumes, respectively. For all registrations, the function `flirt` in FSL (Smith et al., 2004; Jenkinson et al., 2002) was used with search rotation range set between [-90,90] degrees for the three angles and *sinc* interpolation was used for the final interpolation. Using the BET function in FSL (Smith, 2002), a brain mask for each subject was obtained from the axial T1wGd image and applied to all the other registered cMRI and qMRI volumes (for anonymization, but to also make the tumor detection and segmentation tasks easier). The final volumes had a size of 512×512×[24, 36] voxels and a resolution of 0.43×0.43×[4.4, 6] millimeters (mm) in the x, y and z direction, respectively. Intensity normalization was applied to each subject and modality independently, where intensities were scaled into the [-1, 1] range using the 0.5 and 99.5 percentile values as minimum and maximum, respectively. Given the limited number of available subjects, and the low resolution in the z-direction, 2D CNN were used instead of 3D CNN. In total, 528 transversal slices (of which 136 contained tumor) were used for both the tumor detection and segmentation tasks (see section 2.3). Transversal images were used given the higher in-plane resolution. Example images from the dataset are presented in Figure 1.

**Figure 1:**
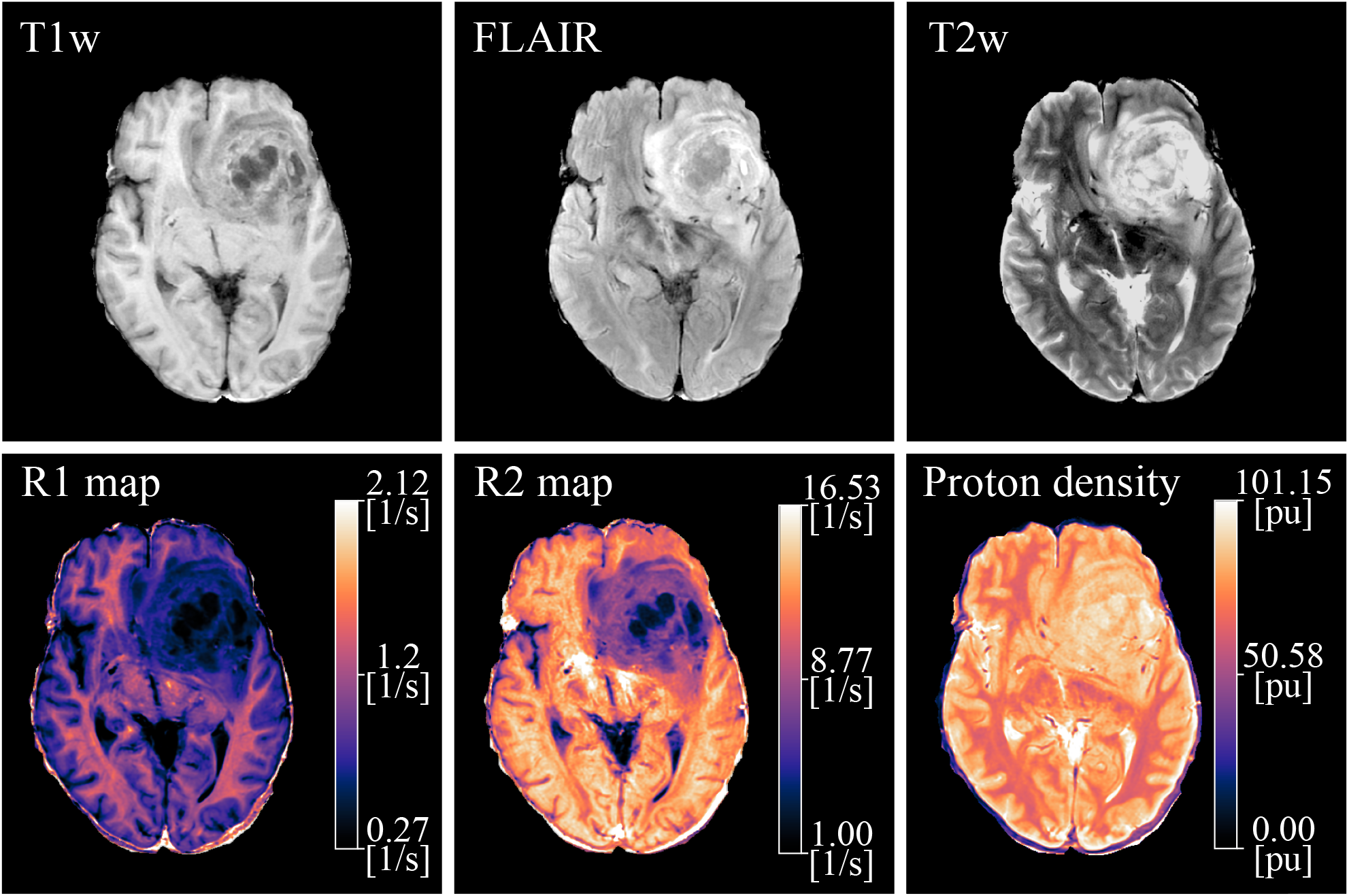
Transversal slice from one subject for six of the MR sequences available. The conventional images (axial T1w, T2w and T2wFLAIR) contain arbitrary values, while R_1_ (1/T1), R_2_ (1/T2) and proton density maps describe quantitative values.

### 2.3. Task definition

To investigate if qMRI data provides additional information compared to cMRI, deep learning models were trained using either conventional or quantitative MRI data as input in the context of tumor detection and segmentation. For the tumor segmentation task, deep learning models were trained to output segmentation masks matching the manual annotations. However, as mentioned earlier, annotations of the tumor core were obtained from T1wGd data thus, they do not account for the non-enhancing infiltrative tumor region invisible in the cMR images, that might be present in the peritumoral edema and captured by the qMRI data. The impact of imperfect annotations, also called weak annotations, on medical image segmentation shows that model training using weak annotations with biased errors, *i*.*e*., errors that consistently alter the annotation, have a negative impact on model performance (Vorontsov and Kadoury, 2021). Thus, evaluating if qMRI provides any additional information compared to cMRI solely on the results of tumor segmentation is not sufficient. A way to disentangle the model training from the weak annotations, while still investigating if qMRI provides additional information compared to cMRI, is to train the models for a simpler task, namely tumor detection. In this study, tumor detection is defined as a binary classification where a deep learning model was trained to classify whether a 2D transversal image of the brain contains a tumor or not. While still using the annotations to label the 2D transversal images with or without tumor, the model was not penalized when using all the information available in the image, thus, limiting the impact of the weak annotation on model training.

### 2.4. Deep learning models

Two different model architectures were used, a shallow 2D CNN classifier for the tumor detection task, and a 2D U-Net model for the segmentation task. For both tasks, data augmentation was used during training by means of random rotation (range=±90 degrees), shift (range=±0.1 of image width), zoom (range=±0.2 of image width), and horizontal and vertical flip. Models were implemented in python using TensorFlow 2.6.0 and training was performed on a workstation with two Nvidia RTX 2080Ti graphics cards with 11 GB of memory each. Training times amounted to a total of 60 days for models trained for tumor detection (four input configurations with a ten-times repeated five-fold cross validation scheme, see section 2.6) and 190 days for the tumor segmentation model (four input configurations with a five-times repeated five-fold cross validation scheme, see section 2.6). For a detailed description of the other packages used in this study, see the code repository available at https://github.com/IulianEmilTampu/qMRI_and_DL.

#### 2.4.1. Tumor detection model (2D-SDM4)

The detection model (hereafter called 2D-SDM4) was a custom-implemented model composed of 4 convolutional blocks functioning as model encoder, followed by two fully connected layers performing the classification. A detailed description of the model architecture can be seen in Figure 2. The model architecture was kept as simple as possible while still allowing for high performance on the classification task. During model training, the binary cross entropy loss between the labels and the model prediction was minimized using the Lookahead optimizer (Zhang et al., 2019) with Adam (Kingma and Ba, 2014) inner optimizer (sinc_period=5, slow_step_size=0.5). The learning rate was set to 1e-6 and kept constant during training. Models were trained for 300 epochs without early stopping.

**Figure 2:**
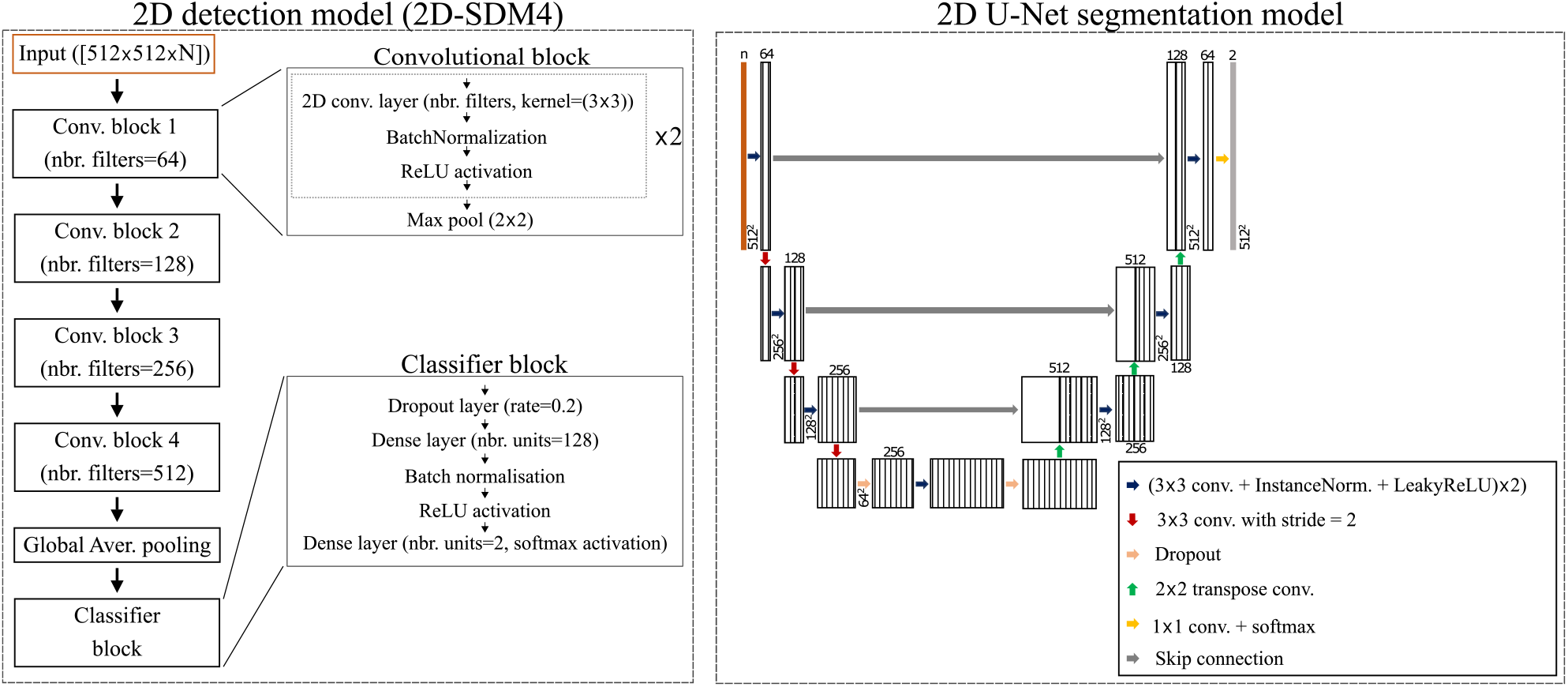
Model architecture summary for the 2D detection (2D-SDM4) and the 2D U-Net segmentation models. The input for both models had size [512×512×N], where N specifies the number of input channels which changed based on the input configuration (4 for cMRI, 3 for qMRI and qMRI_Gd, and 6 for qMRI+qMRI_Gd).

#### 2.4.2. Tumor segmentation model

A 2D U-Net model (Ronneberger et al., 2015) was trained for tumor segmentation given the state of the art performance achieved by this model on a variety of medical image segmentation tasks (Isensee et al., 2021). In this work, the original implementation of U-Net described in (Ronneberger et al., 2015) was adjusted following the description of the architecture template provided by (Isensee et al., 2021). A summary of the model architecture is presented in Figure 2. During training, the sum of weighted Dice score (Sudre et al., 2017) and binary cross entropy loss, as described in (Isensee et al., 2021) was minimized using the Lookahead optimizer (Zhang et al., 2019) with Adam (Kingma and Ba, 2014) inner optimizer (sinc_period=5, slow_step_size=0.5). Weights for the tumor and the background class were used in the computation of the loss to address class imbalance existing between the two classes (weights computed on the training dataset). The model was trained for 1000 epochs without early stopping and the best model according to the validation loss was used for comparison. Initial learning rate was set to 0.001 and reduced during training using a polynomial decrease, as described in (Isensee et al., 2021).

### 2.5. Training, validation and test split

Given that the models were trained on 2D images obtained from volumetric data, a per-subject split strategy was used to avoid biasing the evaluation of model performance due to data leakage (Yagis et al., 2021; Tampu et al., 2022). In particular, for every cross validation repetition (see section 2.6), three subjects (14% of the available data) were randomly selected for testing while the remaining were used for training (72% of the available data) and validation (14% of the available data).

### 2.6. Evaluation metrics and statistical analysis

For both tasks, the models were trained using an *n*-times repeated five-fold cross validation scheme to ensure the reliability of the presented results (Consortium, 2010), and to minimize the effects of selection bias of the test set which can affect the study given the limited amount of available data (Reddy et al., 2010). *n* was set to 10 and 5 for the tumor detection and segmentation tasks, respectively. Tumor detection performance on the test dataset was evaluated in terms of Matthews correlation coefficient (MCC) since it is stable to class imbalance (Chicco and Jurman; Chicco et al., 2021). In addition, precision, recall, accuracy and F1-score were computed using the definition of (Sokolova and Lapalme, 2009), and the receiver operator characteristic (ROC) curve with the corresponding area under the curve (AUC) were also reported. Tumor segmentation performance was instead evaluated in terms of the Dice similarity coefficient (Sudre et al., 2017). The Wilcoxon signed-rank test (two-tailed), was used to compare the models’ performance between models trained using different input configurations. A *p*-value < 0.05 was considered to show a significant difference. Bonferroni correction for multiple comparisons was applied to reduce the risk of type I error.

### 2.7. Model explainability

#### 2.7.1. Occlusion mapping

For the task of brain tumor detection, occlusion mapping (Zeiler and Fergus, 2014) was performed to generate visual descriptions of the brain regions that influenced tumor detection. By analyzing explainability, this study seeks to investigate: (1) what are the regions in the input image that substantially influence model’s detection of the tumor, (2) if there are differences between these regions for models trained on cMRI or qMRI data, and (3) if these differences provide additional information with respect to the nature of the tumor that can help radiologists to improve treatment planning.

Occlusion mapping is a perturbation-based model explainability method through which, for a given image, the relevance of a region towards a class of interest (in this case tumor presence) is measured by how much the model’s predicted probability (*i.e*., softmax score) for the class of interest changes when the region is occluded (*i.e*., its pixels are set to background value: −1 in this study), compared to the model’s prediction on the not-occluded image. Commonly, a squared occlusion mask is moved over the entire image to obtain a spatially resolved occlusion relevance map that shows what regions in the image are important for the classification of the class of interest. The smaller the size of the occlusion mask, the higher the spatial resolution of the relevance map at the cost of a longer computational time. In this study, the occlusion mask size was set to 5×5 pixels to obtain a high resolution relevance map for all test images and all models trained through the repeated cross validation scheme. By taking the average relevance map for each test image over the different models and thresholding it using a value of T=0.03, a larger relevant region was identified. The relevant regions for models trained on different input configurations were computed and compared. A schematic representation of how the relevant regions were obtained from the occlusion relevance maps is shown in Figure 3.

**Figure 3:**
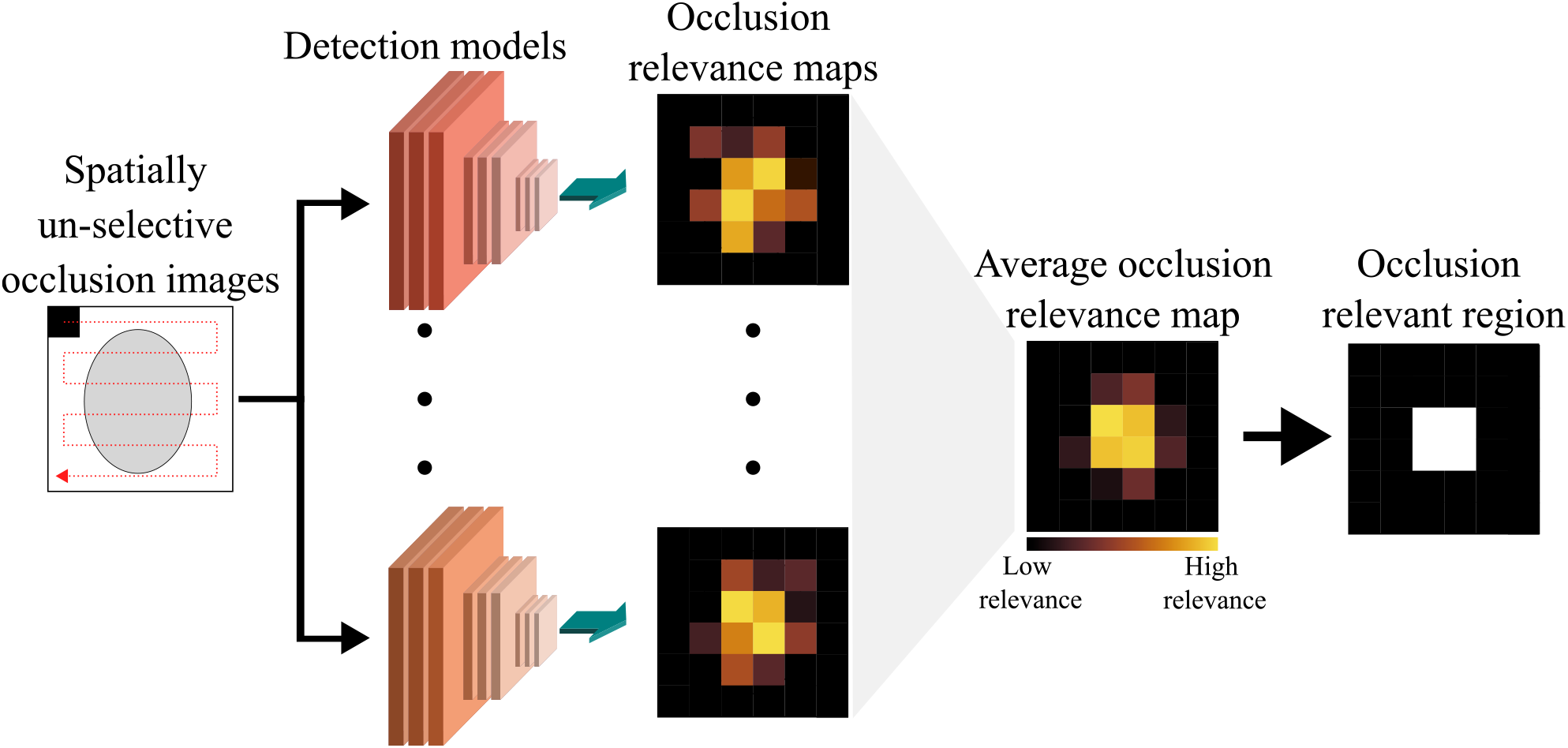
Identification of image regions relevant for tumor detection. The occlusion relevance maps were obtained by performing a spatially non-selective occlusion mapping with a small patch (5×5) using the models trained through the repeated cross validation scheme. The occlusion relevance maps were then averaged and thresholded (T=0.03) to obtain a larger spatially selective occlusion relevant region. The average occlusion relevance map was obtained by averaging all the models from the repeated cross validation scheme which were not trained on a given image.

The importance of the brain region identified by the relevant regions was evaluated by computing the difference between the models’ predicted probability for the tumor presence class on the occluded image (by relevant region) and the not-occluded image. A statistical evaluation of the impact of the relevance region on model prediction was not performed since, to the best of our knowledge, there is no established method to statistically compare predictions on paired images (original and occluded) for models trained through cross validation. Thus, a qualitative comparison between relevant regions identified by models trained on different input configurations was performed by plotting the distribution of R_1_ and R_2_ relaxation ratex (as probability densities) pre- and post-contrast injection, and differentiated between relevant regions falling inside or outside the tumor annotation.

#### 2.7.2. Gradient class activation mapping (GradCAM)

In addition to the occlusion mapping analysis, GradCAM (Selvaraju et al., 2017) was also used to allow visual comparison between the two explainability methods. GradCAM is a feature attribution method that highlights regions in the input image which are relevant when predicting a chosen class (range in [0,1], with 0=irrelevant, 1=most relevant). In this study, GradCAMs were computed on the last convolutional layer of the tumor detection models with respect to the tumor presence class. To quantify how much of the relevant regions fall in the region of visual explanation, the fraction of the GradCAM values that were within the tumor annotation was computed. This approach is similar to that proposed by (Arias-Duart et al., 2022) as a way to quantify model explainability results.

Moreover, the models’ ability to focus on the region of visual explanation identified by the tumor area was investigated using mosaic images. This was initially suggested by (Arias-Duart et al., 2022) as a method to quantify explainability methods, and used here to visualize the ability of the model to focus on the tumor area when presented with unseen images constructed to contain multiple brains.

In particular, mosaic images were obtained by combining in a two-by-two grid four randomly selected images. Of the four images composing the mosaic, two were selected to contain tumor and two not to contain tumor. Moreover, the position of the images in the two-by-two grid was randomly assigned. GradCAM and occlusion mapping were performed on the mosaic images as described above. Note that in the case of the mosaic image, the GradCAMs and the predictions from all the 50 models trained through the repeated cross validation scheme were used since the mosaic images are examples that none of the models has been trained on (model training was performed on transversal slices and not mosaic images).

## 3. Results

### 3.1. Tumor detection

Tumor detection performance on the test data for the models trained on the different input configurations is summarized in Table 1, with results reported as mean±standard deviation over the 50 models trained through ten-times repeated five-fold cross validation scheme. In addition, ROC and box plots for MCC are shown in Figure 4. The model trained on a combination of qMRI data pre- and post-contrast (qMRI+qMRI_Gd) achieved the highest score for all the metrics. However, the difference in performance was significant only when compared to those of the model trained on qMRI pre-contrast data (for all the metrics). No statistical significant difference in model’s performance was found when comparing the models using cMRI, qMRI_Gd or qMRI+qMRI_Gd. It is not clear if this is due to no actual difference, or due to too low statistical power.

**Table 1.** Summary of tumor detection performance for models trained on different input configurations. Results are presented as mean±standard deviation (m±std) over the models trained using the ten-times repeated five-fold cross validation scheme. The model with the highest performance for the different metrics is shown in bold.

| Input configuration | MCC [-1,1]<br>(m±std) | F1-score [0,1]<br>(m±std) | AUC [0,1]<br>(m±std) | Accuracy [0,1]<br>(m±std) | Precision [0,1]<br>(m±std) | Recall [0,1]<br>(m±std) |
| --- | --- | --- | --- | --- | --- | --- |
| cMRI (BraTS) | 0.67±0.15 | 0.81±0.11 | 0.89±0.10 | 0.86±0.07 | 0.90±0.09 | 0.80±0.10 |
| qMRI | 0.49±0.24 | 0.73±0.13 | 0.84±0.09 | 0.80±0.07 | 0.83±0.10 | 0.73±0.12 |
| qMRI_Gd | 0.70±0.15 | 0.84±0.09 | 0.91±0.07 | 0.88±0.05 | 0.91±0.07 | 0.83±0.09 |
| qMRI+qMRI_Gd | **0.72±0.13** | **0.85±0.07** | **0.92±0.06** | **0.88±0.05** | **0.92±0.06** | **0.84±0.08** |

**Figure 4:**
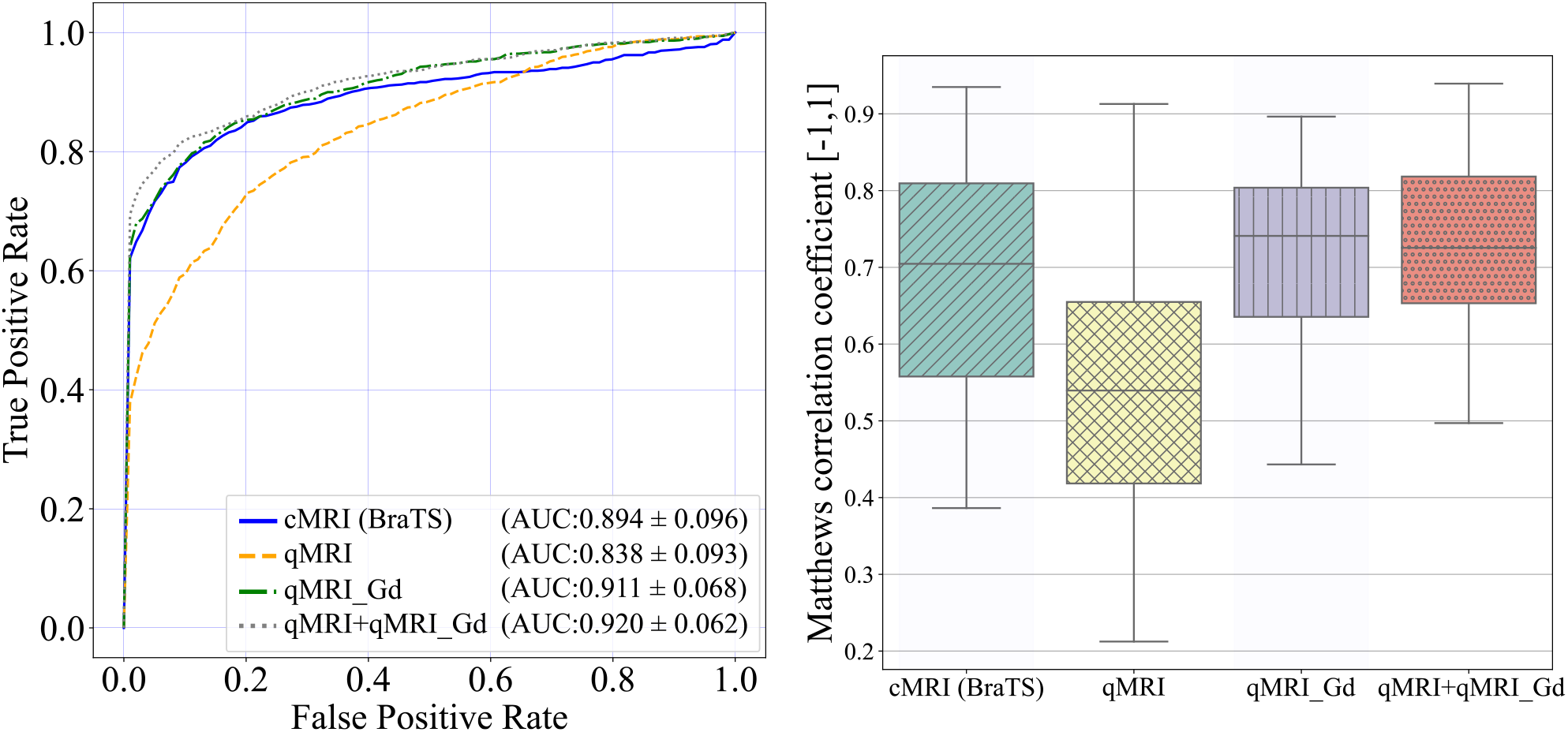
Receiver-operator curves (left) and Matthews correlation coefficient (right) for the models trained on different input configurations for the task of tumor detection. Each box plot summarizes the test performance for the models trained through a ten-times repeated five-fold cross validation scheme.

### 3.2. Model explainability for tumor detection

In the following section, the results of GradCAM and occlusion mapping on mosaic images are presented followed by examples of relevance regions obtained from occlusion mapping on single brain images.

Figure 5 shows examples of GradCAM and occlusion relevance maps performed on mosaic images composed by slices with small (Figure 5.a) and large (Figure 5.b) tumor sizes, and for the models trained for tumor detection. The regions highlighted as important for tumor detection are coherent between the two explainability methods. The higher spatial resolution of the occlusion maps shows even smaller regions which are lost in the GradCAMs (see small tumor cases). Overall, the regions highlighted as important for the tumor detection are larger than the manual annotation and models trained on input configurations that included qMRI post-contrast data showed to localize the tumor region in most of the cases, whereas models using cMRI or qMRI pre-contrast did not focus on the tumor area in all the instances where tumor was present. These results, obtained on constructed images that the models have not been trained on, show the ability of the models to focus on regions of visual explanation when performing classification. Table 2 summarizes the quantification of GradCAMs relevance maps on the original dataset images (single brain transversal slices). Overall, no trends could be seen with respect to the different input configurations.

**Table 2.**
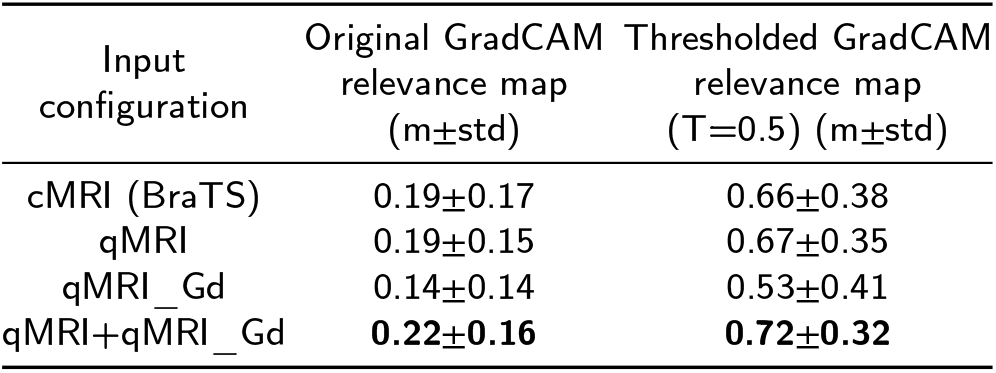
Summary of the fraction of relevance regions identified by GradCAM falling in the region of visual explanation, *i.e*. the manual annotation of the tumor structure (higher values = high overlap). Values are computed over single transversal images, and are presented for the original and thresholded (T=0.5) GradCAM relevance maps, and for each input configuration as mean±standard deviation (m±std) over the transversal slices showing tumor structure (n=136). The input configuration that achieved the highest overlap between Grad-CAM and tumor annotation is in bold.

| Input configuration | Original GradCAM relevance map<br>(m±std) | Thresholded GradCAM relevance map<br>($T=0.5$) (m±std) |
| --- | --- | --- |
| cMRI (BraTS) | 0.19±0.17 | 0.66±0.38 |
| qMRI | 0.19±0.15 | 0.67±0.35 |
| qMRI_Gd | 0.14±0.14 | 0.53±0.41 |
| qMRI+qMRI_Gd | **0.22±0.16** | **0.72±0.32** |

**Figure 5:**
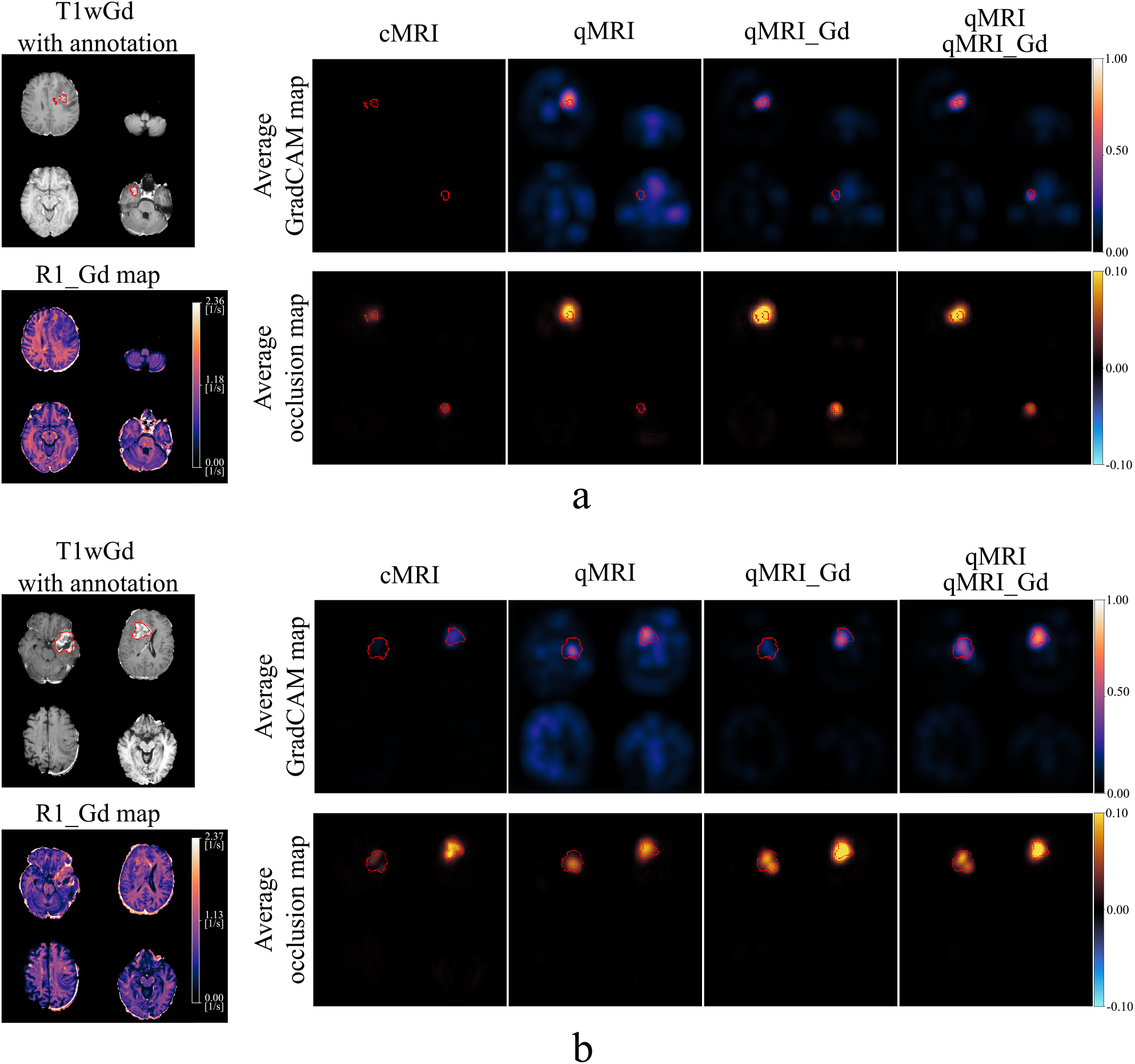
Examples of model explainability analysis on mosaic images for models trained for tumor detection with different input configurations. For each example (a and b), the T1wGd mosaic image is shown with the manual annotation contour overlaid in red (left-most column). Moreover, for each input configuration and example, the GradCAM (top row) and occlusion (bottom row) relevance maps are presented with the tumor annotation contour overlaid in red. GradCAM and occlusion relevance maps were computed with respect to the positive class (tumor presence) and are shown as the mean over the 50 models trained through the ten-times repeated five-fold cross validation. Image best viewed in colors.

Examples of GradCAM and occlusion mapping analysis for transversal slices are presented in Figure 6. GradCAM and occlusion relevance maps highlighted similar regions as important for tumor detection. The relevant region, obtained by thresholding the relevance map (T=0.03) when using a 5×5 patch, size is also shown. For both examples, the predicted probability for the tumor presence class was substantially reduced (reduction higher than 0.6) when occluding the relevant region for models using cMRI, qMRI_Gd or qMRI+qMRI_Gd input configurations. On the other hand, a reduction smaller than 0.2 in the predicted probability for the tumor class was observed for the models trained on qMRI pre-contrast data. These findings, in combination with the tumor detection classification results, confirm the importance of the contrast agent in highlighting the tumor region and show that the detection models exploit the relation between pre- and post-contrast images.

**Figure 6:**
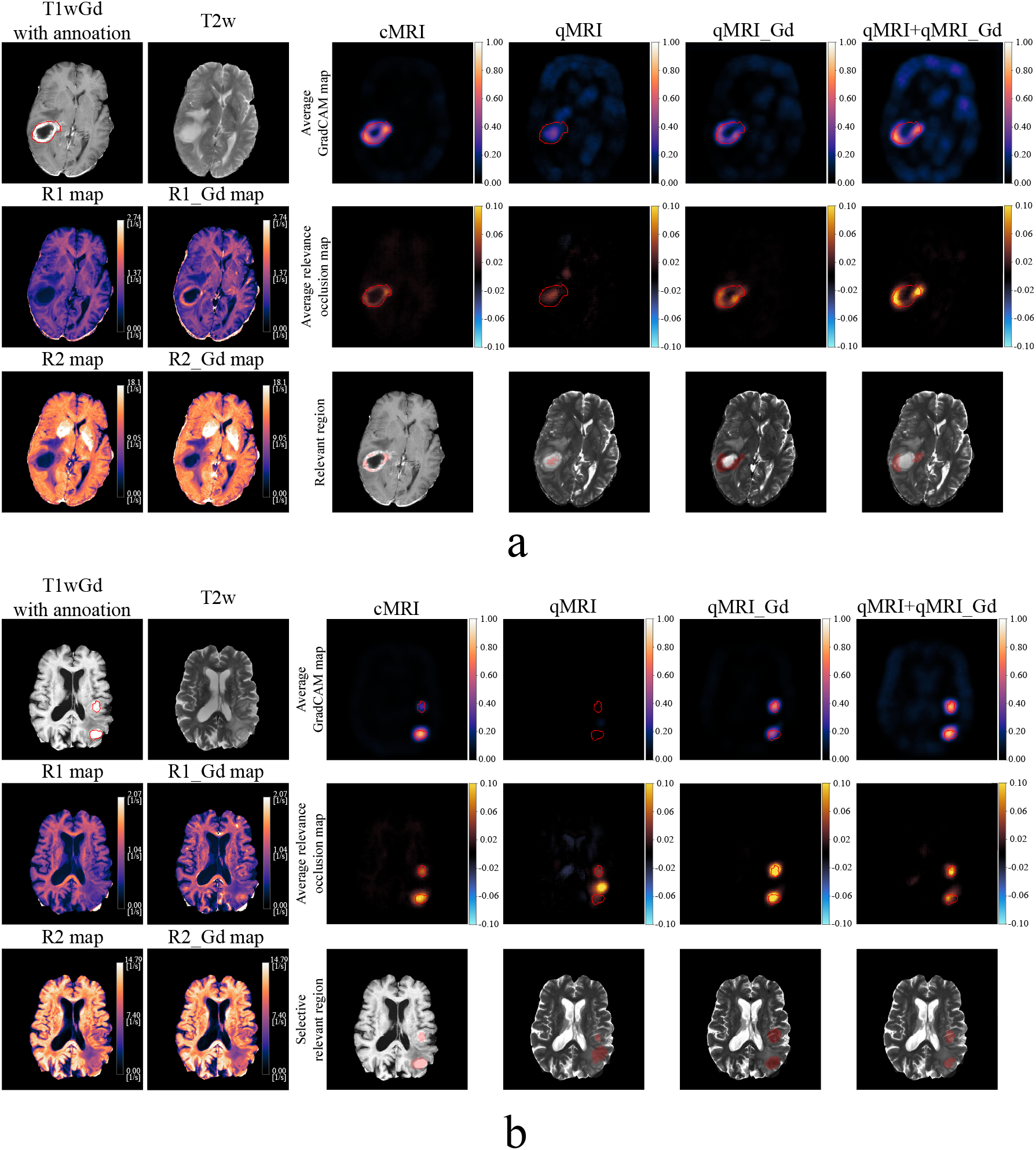
Examples of model explainability analysis on transversal images for models trained for tumor detection with different input configurations. For each example (a and b), the T1wGd image is shown with the manual annotation contour overlaid in red along with the T2w image and the R_1_ and R_2_ pre- and post-contrast quantitative maps. For each example and input configuration the following are presented: (top row - *Average GradCAM map*) the average GradCAM images, (middle row - *Average relevance occlusion map*) the average relevance occlusion map, and (bottom row - *Relevant region*) relevance occlusion mask in red obtained by thresholding the average relevance maps (T=0.03). The average GradCAM and occlusion relevance maps were computed with respect to the positive class (tumor presence) and are shown as the mean over the models which did not use such images during model training. Image best viewed in colors.

Considering models trained on cMRI or qMRI+qMRI_Gd data, the regions identified as relevant for the tumor detection were proximal to or overlapping with the tumor annotation for most of the images in the database (also confirmed by the analysis of the GradCAM in Table 2). This shows that the detection models could focus on the region of visual explanation and exploit this information for tumor detection even though trained only for a binary classification between slices containing or not containing tumor.

By visualizing the R_1_ and R_2_ relaxation rates distribution and shift after contrast injection of the identified relevant regions for all the transversal slices in the dataset having tumor annotation (n=136) and for models trained on either cMRI or qMRI+qMRI_Gd data it can be seen that (1) there is small to no difference in the relaxation rates distribution and shift after contrast injection between relevant regions obtain from models trained on cMRI or qMRI+qMRI_Gd data, and (2) the relevant regions inside the annotation have in increase relaxation rates after contrast injection, while the regions outside do not (see Figure 7). However, these results presented over the entire dataset do not capture the diversity in the glioblastoma tumors available in the dataset and do not account for the differences in relaxation rates between subjects. In fact when performing a similar analysis for each subject independently, different trends can be seen. In the example presented in Figure 8, the relaxation rates of relevant regions both inside and outside the tumor annotation increase after contrast injection, especially for the region identified as relevant by both models trained on cMRI and qMRI data. The change in relaxation rates can be attributed to the deposition of gadolinium in the tissue resulting from the damage of the blood vessel walls caused by the active tumor tissue growth. It is also interesting to note that the R_1_ relaxation for the relevant region inside the annotation (range in [0.24, 0.96]) spread out after contrast injection showing the inhomogeneity of tissue properties in this region (range [0.24, 2.62]). This is also visible for R_2_ relaxation, where the rates’ distribution shows two predominant tissue types (two distinct peaks in the R_2_ relaxation probability density graphs in the violin plots) of which only one has increased R_2_ relaxation rate after contrast injection. Moreover, the shift in relaxation value for the regions inside and outside the annotation follows a similar trend, as can be seen from the violin plot of the pixel-wise difference. This suggests that tissues with similar biological activity is present in these two regions. Additional examples showing a comparison between relevant regions inside and outside the tumor annotation for individual subjects are presented in Appendix B, where Figure 11 shows both a shift and a spread of relaxation rates after contrast injection, and Figure 12 only a shift.

**Figure 7:**
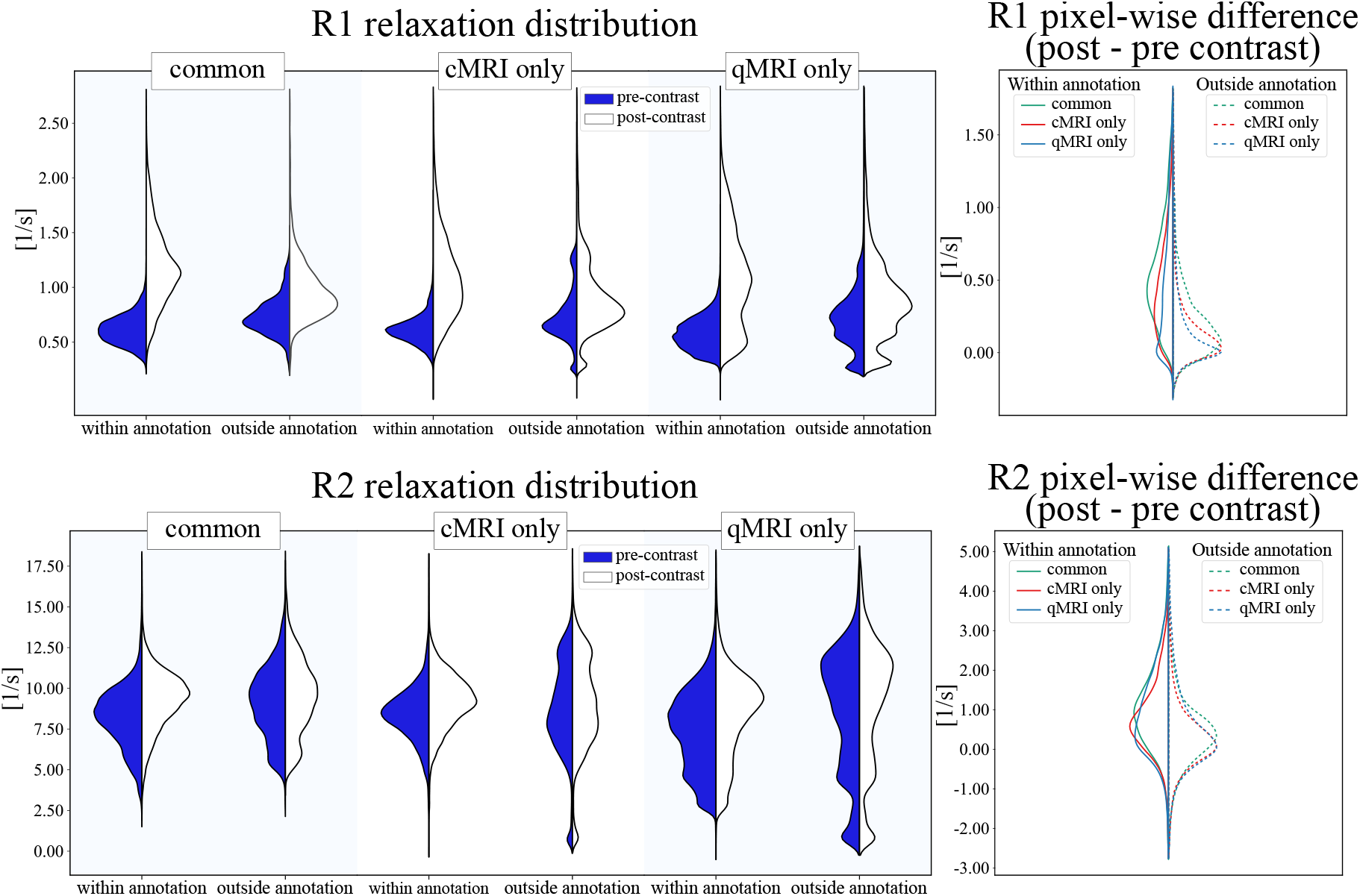
Qualitative comparison of R_1_ and R_2_ relaxation rates of relevant regions obtained from models trained on cMRI and qMRI+qMRI_Gd data over all the transversal slices containing annotated tumor (n=136). R_1_ and R_2_ relaxation rates pre- and post-contrast for the relevant regions within and outside the annotation are shown as violin plots. Quantitative pixel-wise difference pre- and post-contrast for the relevant regions inside and outside the annotation are also shown as violin plots. In the difference graph, the common region (green), the region identified only by cMRI (red), the region identified only by qMRI+qMRI_Gd (blue) are presented with solid and dashed lines within and outside the annotation regions, respectively. Image best viewed in colors.

**Figure 8:**
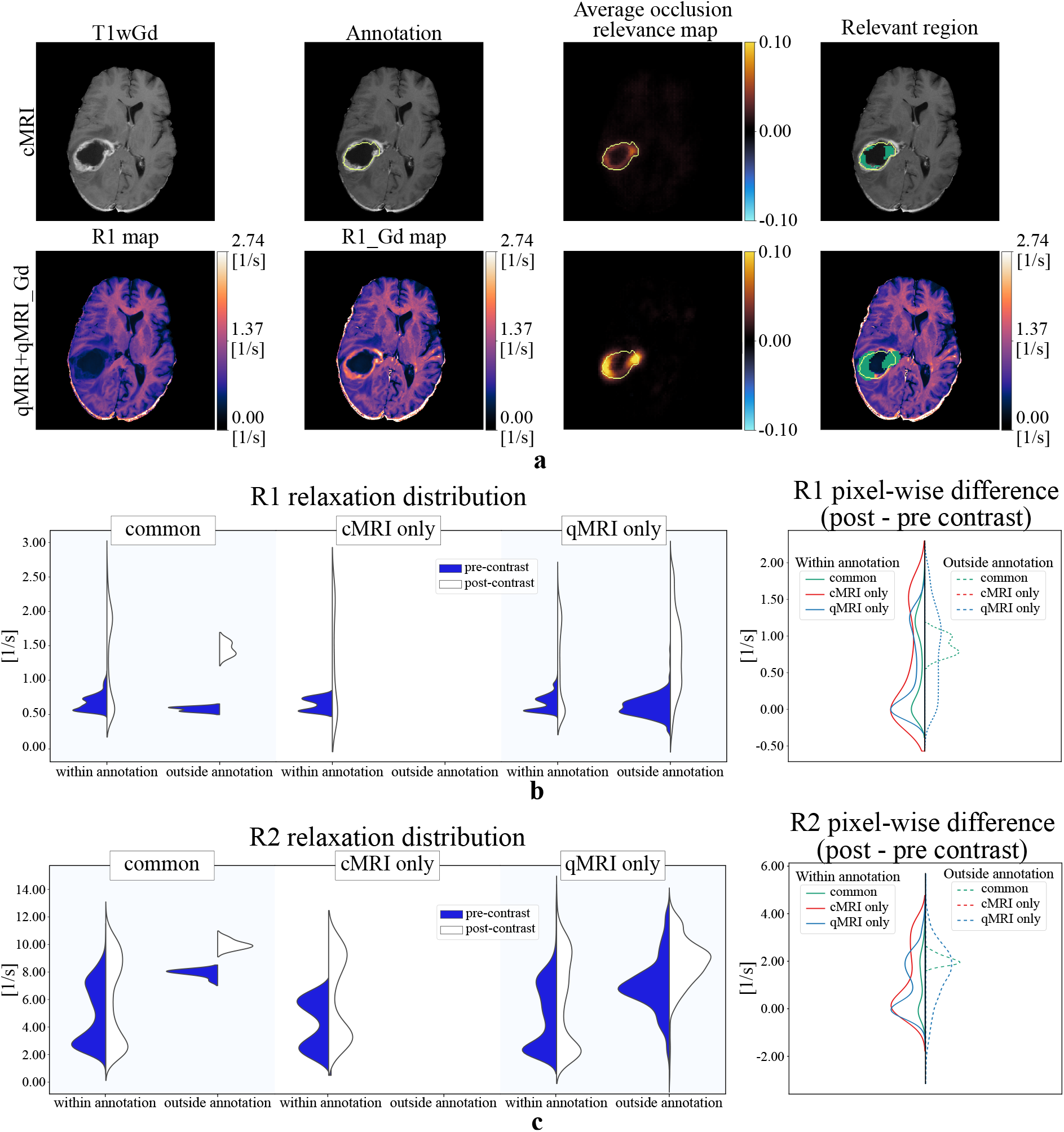
Qualitative comparison of R_1_ and R_2_ relaxation rates of relevant regions obtained from models trained on cMRI and qMRI+qMRI_Gd data. (a) transversal slice as seen with T1wGd and by quantitative R_1_ (pre- and post-contrast). The average occlusion relevance map used to obtain the relevant region is presented with the tumor annotation boundary shown in yellow. The relevant region is presented in green overlayed on T1wGd and R_1__Gd map. R_1_ and R_2_ relaxation rates pre- and post-contrast for the relevant regions within and outside the annotation are shown as violin plots (b and c, respectively). Quantitative pixel-wise difference pre- and post-contrast for the relevant regions inside and outside the annotation are also shown as violin plots. In the difference graph, the common region (green), the region identified only by cMRI (red), the region identified only by qMRI+qMRI_Gd (blue) are presented with solid and dashed lines within and outside the annotation regions, respectively. Image best viewed in colors.

### 3.3. Tumor segmentation

Dice similarity coefficient computed on the test data for the models trained with different input configurations is summarized in Table 3, with results reported as mean±standard deviation over the 25 models trained through five-times repeated five-fold cross validation. Ensemble performance obtained by averaging models’ softmax predictions on the test cases is also presented. In addition, boxplots for DSC are shown in Figure 9. The highest DSC (0.89±0.03) was obtained by the model trained on the qMRI post-contrast data. A significant difference in model performance could only be found when comparing qMRI pre-contrast with any of the other input configurations (*p*<0.008). Representative images of the models’ segmentations when trained on different input configurations are shown in Figure 10. Overall, models’ segmentation were coherent with the manual annotations as summarized by the DSC metric, and no trends could be seen when considering over or under-segmentation for any of the input configurations. Nevertheless, it is interesting to note how the three models using qMRI data over-segmented the region between the two manually annotated tumor regions, while the model trained on cMRI data did not (Figure 10.b). This may suggest that tumor-like features extracted from the quantitative data are present in this region, and that qMRI could perform better than cMRI if using larger annotations.

**Table 3.** Segmentation performance for the models trained with different input configurations. Values are presented as mean±standard deviation (m±std) Dice similarity coefficient (DSC) over the models trained through a five-times repeated five-fold cross validation scheme. Model ensemble performance is also shown. The input configuration achieving the highest DSC is shown in bold. Statistically significant differences could only be found when comparing qMRI with any of the other input configurations.

| Input configuration | DSC (m±std) | ensemble DSC (m±std) |
| --- | --- | --- |
| cMRI (BraTS) | $0.880\pm0.038$ | $0.897\pm0.020$ |
| qMRI | $0.760\pm0.043$ | $0.782\pm0.023$ |
| qMRI_Gd | $0.885\pm0.028$ | $0.906\pm0.011$ |
| qMRI+qMRI_Gd | **$0.883\pm0.027$** | **$0.902\pm0.015$** |

**Figure 9:**
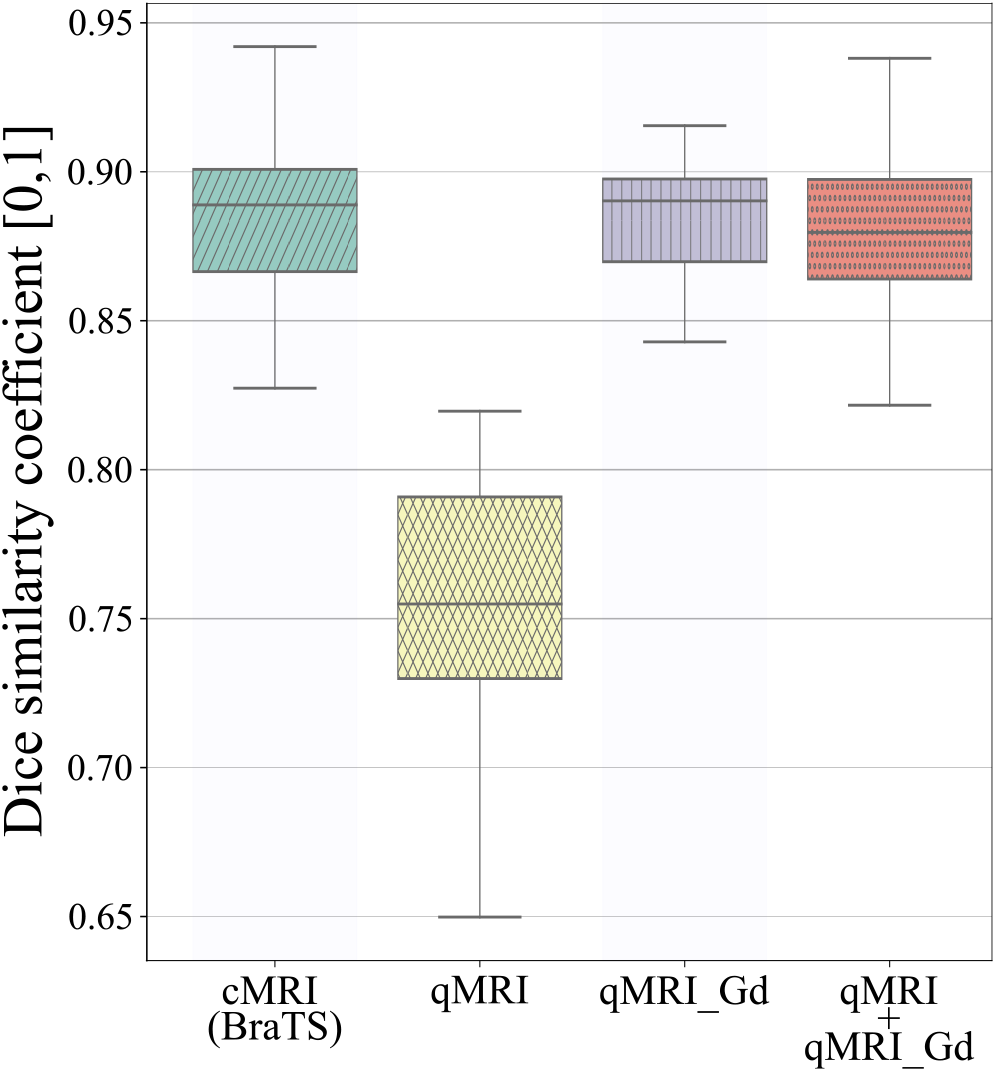
Dice similarity coefficient for the models trained on different input configurations for the task of tumor segmentation. Each boxplot summarizes the test performance for the 25 models trained through a five-times repeated five-fold cross validation. The ground truth annotations were obtained from cMRI data.

**Figure 10:**
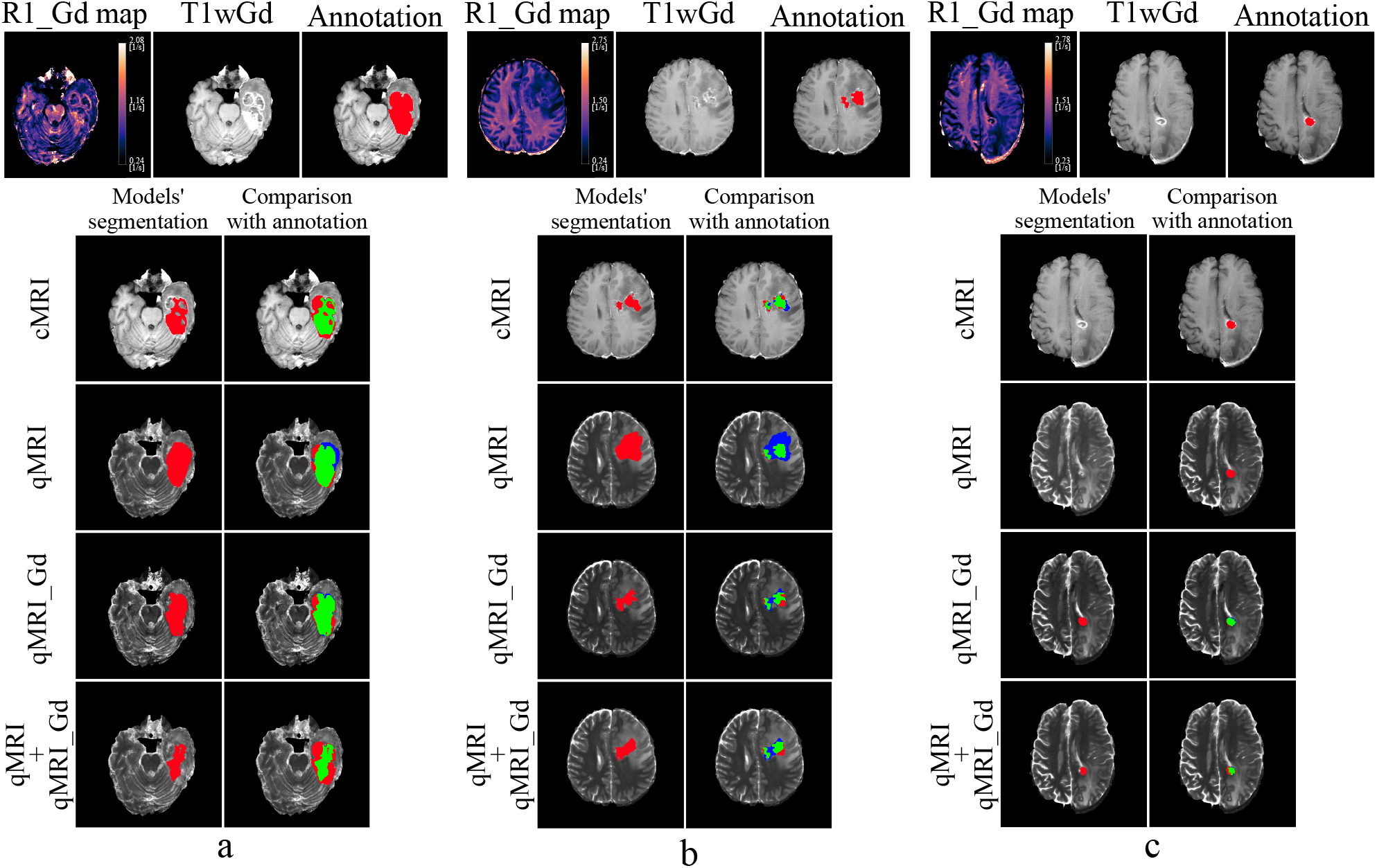
Representative segmentation results for models trained on different input configurations. Quantitative R_1_ map, anatomical T1wGd and the manual annotation (red) overlaid on the T1wGd image are presented on the top row for each example. Model segmentation is presented in two ways: (1) overlayed in red on the anatomical image (left column) and (2) compared to the manual annotation (right column) where green shows true positive, blue false positives (over-segmentation) and red false negatives (under-segmentation). Image best viewed in colors.

## 4. Discussion

In this study, deep learning models for tumor detection and segmentation were trained using either cMRI or qMRI data. Using model explainability methods, relevant regions for the detection of the tumor were identified and the trends in quantitative values pre- and post-contrast in those regions were investigated. We show that (1) tumor detection and segmentation performance for models trained on qMRI_Gd data are on par with those trained on cMRI data, (2) regions in the brain relevant for the tumor detection obtained from the analysis of deep learning explainability maps are proximal or overlap the manual annotation of the tumor structure, and (3) relaxation rates of these regions increase after contrast injection, indicative of the presence of active tumor tissue.

### 4.1. Tumor detection and segmentation

Even though training models on qMRI data did not result in significant improvement in tumor detection and segmentation, obtaining performances on par with cMRI (BraTS) data shows the potential of qMRI as scanner-independent input for deep learning models. In fact, qMRI measures properties of brain tissue properties which do not change depending on MR scanner brand or the field strength. Thus, collecting qMRI data at many different sites (similar to BraTS) would potentially not require any harmonization of the images (which deep learning models normally are sensitive to). In addition, the tissue relaxation properties measured by qMRI can be used post-acquisition to generate synthetic MR images with various contrast-weighing along with automatic tissue segmentation of white matter, gray matter, cerebrospinal fluid and myelin (Gonçalves et al., 2018). It is worth noting that although the tumor detection task was designed to alleviate the impact of the weak annotations on the model training, the presence or not of a tumor structure was still based on the cMRI data. This could have biased the model in only detecting tumor in those slices where tumor is visible in cMRI, disregarding those slices where tumor could only be detected using the qMRI data.

The model performance for both tumor detection and segmentation was significantly lower for models trained on qMRI pre-contrast, compared to all other input configurations. The reason for this is most likely due to the missing hyperintense image regions indicative of active tumor visible after contrast injection. Moreover, the reason why qMRI pre-contrast performs worse than cMRI is probably due to cMRI containing images with and without gadolinium contrast (T1w, T1wGd), and the network can learn to look at the difference between these images.

Specific to the tumor segmentation task, using ground truth annotations obtained on the same input configurations as that used to train the models could show different model performances, especially for those models trained on qMRI data. In fact, all models were trained using as ground truth the annotation of the tumor border obtained from a high resolution T1wGd image, disregarding the input configuration. This could have negatively affected models using qMRI data since the labels the models were trained to match, did not account for all the information available in such data.

### 4.2. Model explainability analysis

No difference in the pre- and post-contrast injection relaxation rate trends could be seen over the entire dataset for the relevant regions identified by models trained on cMRI or qMRI data using the model explainability analysis implemented in this study. Thus, no radiological biomarkers sensitive to tumor detection specific for qMRI could be identified. Nevertheless, from the analysis of individual subjects it could be seen that relaxation rate changes after contrast injection for relevant regions outside and proximal to the tumor annotation were similar to those of regions within the annotation, indicative of the presence of tumor-like tissue proximal to the visible tumor area. These findings are coherent with those of (Blystad et al., 2017) in showing contrast-enhancing induced changes in relaxation rates in the peritumoral edema. Moreover, the spread in relaxation rates after contrast injection seen for some of the examples for relevant regions outside the tumor annotation is indicative of a high inhomogeneity in tissue properties in those areas. In the context of glioblastoma, this inhomogeneity can be attributed to the concomitant presence of vasogenic edema and infiltrative tumor (Hattingen et al., 2013; Oh et al., 2005) in the edema region proximal to the contrast-enhancing tumor. Further investigation through image guided biopsies is needed to validate such regions.

### 4.3. Limitations

The small number of subjects available in the dataset (n=21) hinders the generalizability of the findings of this study, especially when looking at the comparison between the regions identified by the models to be useful for tumor detection when using cMRI or qMRI data. A larger number of subjects would increase the statistical power to provide stronger evidence for the trends in quantitative values of the regions identified as important for the tumor detection. More subjects would also make it possible to train deep learning models on 3D volumes instead of 2D slices, using 3D CNNs. Moreover, by also including a larger variety of tumor types and grades, the presented approach for the identification of regions important for tumor detection could highlight differences between tumor types and grades which could be used during diagnosis.

The in-plane resolution is high (0.43×0.43 mm), but the slice thickness ranging from 4.4 to 6 mm results in the quantitative values in each pixel to reflect a variety of tissues. This is exacerbated in those regions in the brain where there is transition between tissue types, such as at the border of the tumor. For this reason, the analysis in this study was limited in only showing overall trends in quantitative values of the relevant regions identified by the models. Moreover, the pre-processing needed to align the manual annotation of the tumor border to the qMRI data introduced interpolation artefacts which hindered any conclusion from the analysis of the quantitative value trends for the small tumor regions.

## 5. Conclusion

In this study, the potentially added information from qMRI data in the context of tumor segmentation and detection was investigated. Analysis of quantitative relaxation rates of brain regions identified relevant for tumor detection by model explainability maps was also performed. Models trained on qMRI post contrast data show comparably high performance in tumor detection and segmentation as models trained on cMRI data, with the advantage of qMRI measuring tissue relaxation, potentially removing the need for data harmonization and possibility of generating synthetic contrast-weighted MR images. No distinct trends in the relaxation rates between regions identified as relevant for tumor detection by models trained on either cMRI or qMRI data could be seen when considering the entire dataset. On the other hand, the analysis of individual subjects highlighted cases where the model-identified relevant regions outside the tumor annotation showed changes in relaxation rates after contrast injection similar to regions within the visible tumor area, suggestive of infiltrative tumor in the peritumoral edema which are detectable by deep-learning but invisible to the eye. A larger number of subjects and tumor types are however needed to generalize the tumor detection and segmentation results, along with image guided histological examinations to validate the identified relevant regions.

## Data Availability

The data used for this study is not shared. The used code is available at https://github.com/IulianEmilTampu/qMRI_and_DL

## Acknowledgments

This study was supported by CENIIT at Linköping University, ITEA3 / VINNOVA funded project “Intelligence based iMprovement of Personalized treatment And Clinical workflow supporT” (IMPACT), LiU Cancer at Linköping University, the Analytic Imaging Diagnostics Arena (AIDA), the ITEA4 / VINNOVA funded project “Automation, Surgery Support and Intuitive 3D visualization to optimize workflow in IGT SysTems” (ASSIST) (2021-01954), the Åke Wiberg foundation (M22-0088), Medical Research Council of Southeast Sweden (FORSS-234551), and Swedish Research Council (2018-05250).

## A. Description of data acquisition protocol

*Isotropic high resolution BRAVO volume (3D-FSPGR GD)*: axial, FOV 240 × 240 mm, 172 slices, voxel size 0.94×0.94×1 mm, TE=3.2 ms, TR=8.2 ms, TI=450 ms.

*Axial T1w spin echo before and after (T1wGd) contrast agent injection*: axial, FOV 220 × 165, 24 slices, voxel size 0.43 × 0.43 × 5 mm (gap 1 mm), TE = 17.7 ms, TR = 2 524 ms, TI (inversion time) = 798 ms.

*Axial T2w spin echo PROPELLER*: axial, FOV 220 × 220, 24 slices, voxel size 0.43 × 0.43 × 5 mm (gap 1 mm), TE = 95-97 ms, TR = 3000 ms.

*qMRI MAGIC before and after contrast agent injection*: axial, FOV 220 × 180, 24 slices, voxel size 0.43 × 0.43 × 5 mm (gap 1 mm). In total 8 images per slice were measured with TE = 22 ms or 95 ms, TR = 4 000 ms, TI = 170, 670, 1840 or 3840 ms.

## B. Additional examples of relevant region comparison

**Figure 11:**
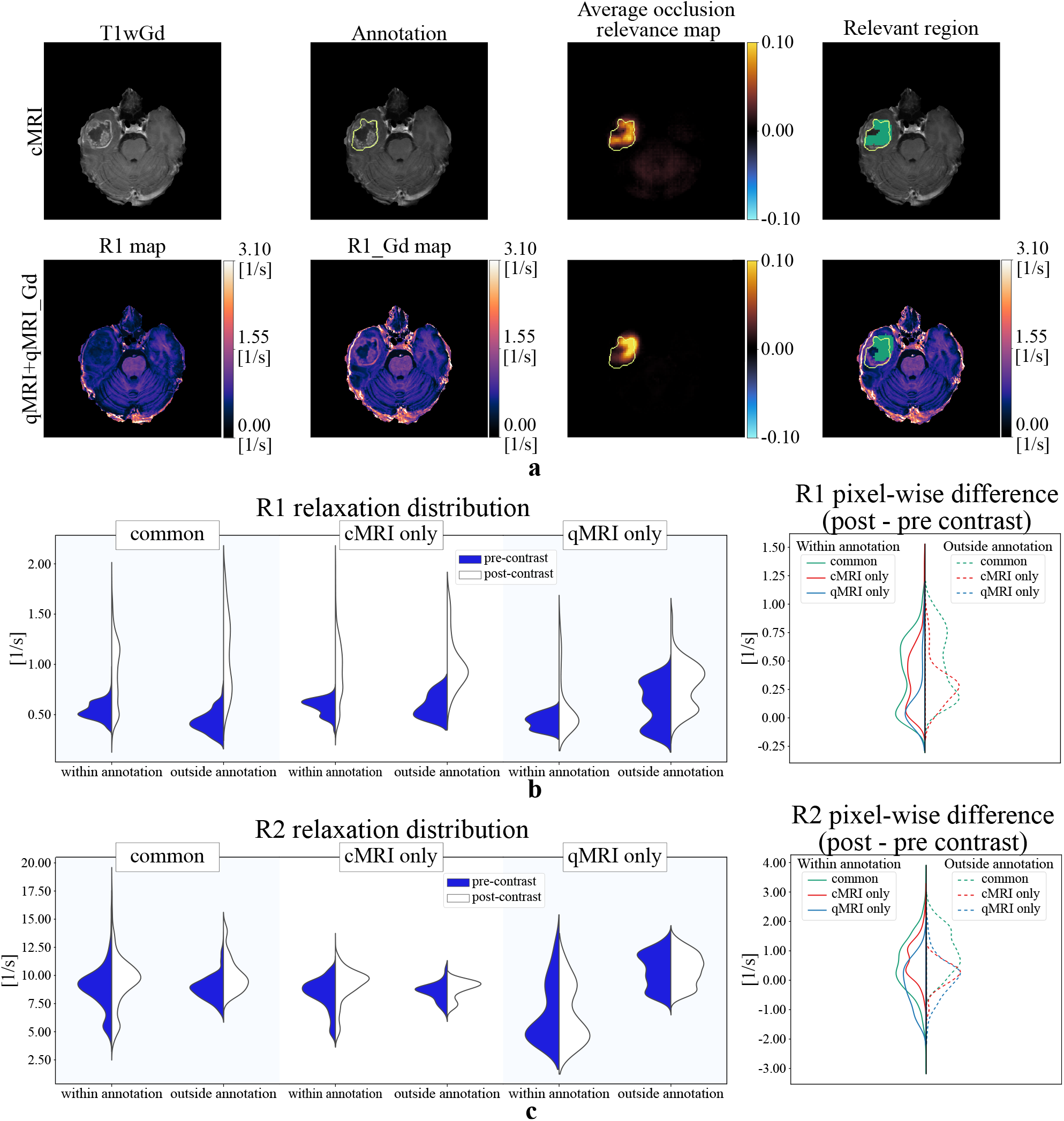
Qualitative comparison of R_1_ and R_2_ relaxation rates of relevant regions obtained from models trained on cMRI and qMRI+qMRI_Gd data. (a) transversal slice as seen with T1wGd and by quantitative R_1_ (pre- and post-contrast). The average occlusion relevance map used to obtain the relevant region is presented, with the tumor annotation boundary shown in yellow. The relevant region is presented in green overlayed on T1wGd and R_1__Gd map. R_1_ and R_2_ relaxation rates pre- and post-contrast for the relevant regions within and outside the annotation are shown as violin plots (b and c, respectively). Quantitative pixel-wise difference pre- and post-contrast for the relevant regions inside and outside the annotation are also shown as violin plots. In the difference graph, the common region (green), the region identified only by cMRI (red), the region identified only by qMRI+qMRI_Gd (blue) are presented with solid and dashed lines within and outside the annotation regions, respectively. Image best viewed in colors.

**Figure 12:**
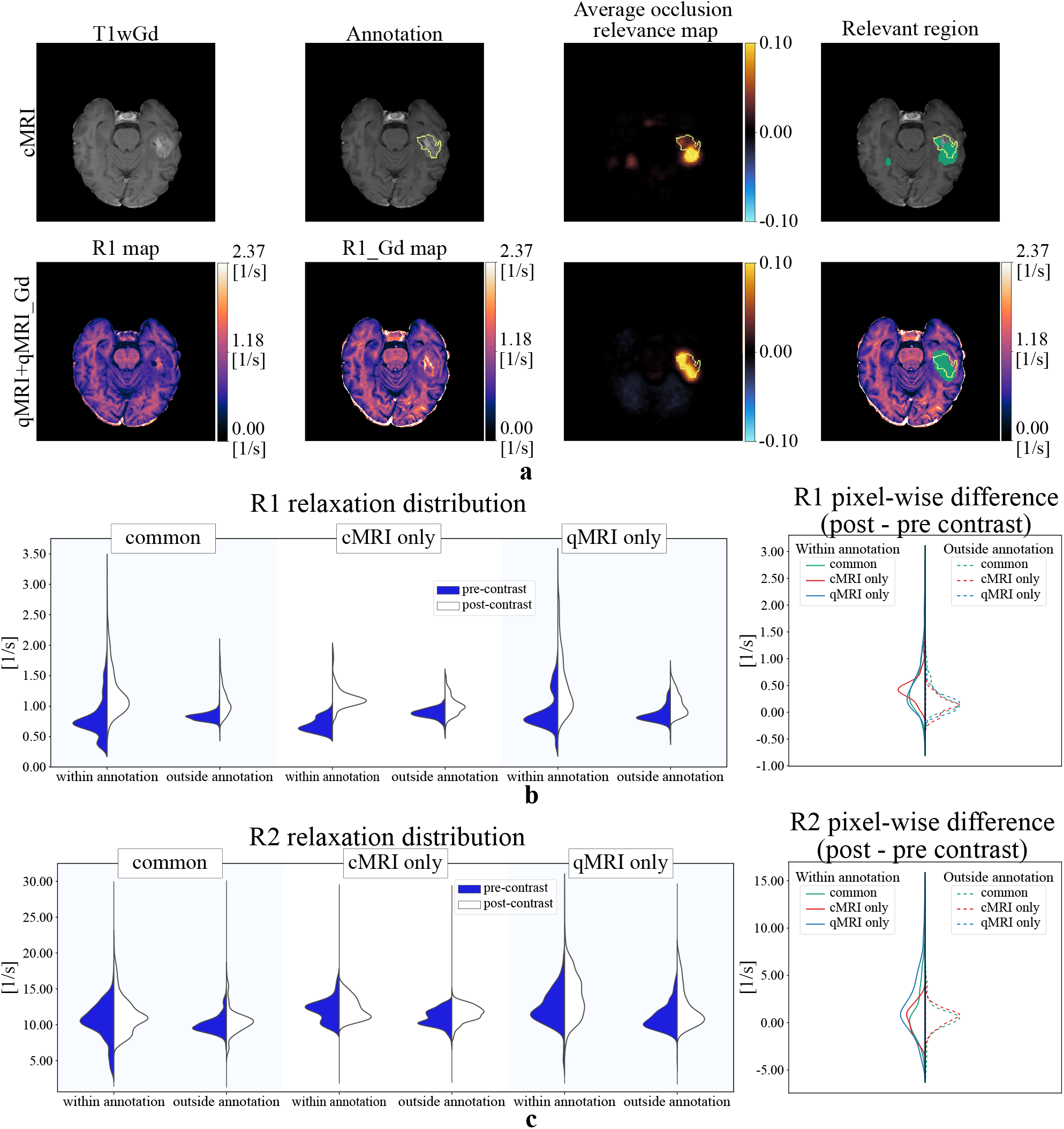
Qualitative comparison of R_1_ and R_2_ relaxation rates of relevant regions obtained from models trained on cMRI and qMRI+qMRI_Gd data. (a) transversal slice as seen with T1wGd and by quantitative R_1_ (pre- and post-contrast). The average occlusion relevance map used to obtain the relevant region is presented, with the tumor annotation boundary shown in yellow. The relevant region is presented in green overlayed on T1wGd and R_1__Gd map. R_1_ and R_2_ relaxation rates pre- and post-contrast for the relevant regions within and outside the annotation are shown as violin plots (b and c, respectively). Quantitative pixel-wise difference pre- and post-contrast for the relevant regions inside and outside the annotation are also shown as violin plots. In the difference graph, the common region (green), the region identified only by cMRI (red), the region identified only by qMRI+qMRI_Gd (blue) are presented with solid and dashed lines within and outside the annotation regions, respectively. Image best viewed in colors.

## Notes

### Competing Interest Statement

The authors have declared no competing interest.

### Funding Statement

This study was supported by CENIIT at Linkoping University, ITEA3 / VINNOVA funded project "Intelligence based iMprovement of Personalized treatment And Clinical workflow supporT" (IMPACT), LiU Cancer at Linkoping University, the Analytic Imaging Diagnostics Arena (AIDA), the ITEA4 / VINNOVA funded project "Automation, Surgery Support and Intuitive 3D visualization to optimize workflow in IGT SysTems" (ASSIST) (2021-01954), the Ake Wiberg foundation (M22-0088), Medical
Research Council of Southeast Sweden (FORSS-234551), and Swedish Research Council (2018-05250).

### Author Declarations

Regional ethical board of Linkoping, Sweden (decision number 2011 / 406-31)

